# Comparing the effectiveness of secondary distribution of HIV self-testing to testing card referral in promoting HIV testing among men who have sex with men in China: A quasi-experimental study

**DOI:** 10.1101/2021.05.09.21256226

**Authors:** Yongjie Sha, Yuan Xiong, Yehua Wang, Jason Ong, Yuxin Ni, Ying Lu, Mengyuan Cheng, Joseph Tucker, Weiming Tang

## Abstract

**Background:** Social network-based HIV self-testing (HIVST) is useful to promote HIV testing. Secondary distribution is one social network-based method whereby individuals (indexes) access multiple HIVST kits and distribute them to their social networks (alters). This quasi-experimental study compared the effectiveness and cost of two social network-based HIV testing strategies (HIVST secondary distribution and HIV testing card referral) in promoting HIV testing among Chinese men who have sex with men (MSM).

**Methods:** MSM aged 18 years or older were recruited in Guangzhou, Guangdong Province. From May to September 2019, indexes recruited during that period could distribute HIVST kits to people within their social network. Indexes recruited from October 2019 to January 2020 could refer HIV testing cards to people within their social network for free facility-based tests. Participants could access 1-5 HIVST kits or testing referral cards for distribution. Alters were encouraged to upload a picture of their test results and complete an online survey. Indexes and alters received an incentive to report test results.

**Results:** Two hundred thirty-nine potential participants were assessed for eligibility and 208/245 (84.9%) were eligible. Among those who completed baseline assessment, 154/208 (74.0%) completed one month of follow-up. Overall,106 indexes were recruited in the HIVST arm and 102 in the testing card arm. The two arms had similar socio-demographic characteristics. At the one-month follow-up, 92 indexes in the HIVST arm self-reported having distributed self-test kits to 179 unique alters, and 62 in the testing card arm self-reported having distributed testing referral cards to 26 unique alters. Additionally, 69/92 (75%) in the HIVST arm distributed any test to friends or sexual partners compared to 18/62 (29%) in the testing card arm, with a risk difference of 46% (95% CI 31%, 61%). Indexes in the HIVST arm distributed an average of 1.95 (SD=1.90) tests, compared to 0.42 (SD=0.78) in the testing card arm, with a risk difference of 1.53 (95% CI 1.09, 1.96). Subgroup analysis suggested that indexes in the HIVST arm who self-identified as gay (p = 0.007) or were previously tested for HIV (p = 0.02) were more likely to distribute. The HIVST arm had a higher total cost and higher testing coverage compared to the testing card referral arm. The ICER per alter tested was $52.78.

**Conclusions:** Secondary distribution of HIVST engaged more MSM to distribute tests to their social network and reached more MSM for test. MSM who self-identify as gay or who have previously tested for HIV were more effective in distributing tests. Future testing approaches should include HIVST kits in voluntary counselling and testing settings and incorporate digital strategies for secondary distribution.

## Background

Men who have sex with men (MSM) accounted for 25.5% of new HIV infections in China(1). HIV testing facilitates early diagnosis and presents a crucial entry point into the HIV care continuum. However, approximately 30% of people living with HIV in China are unaware of their serostatus(2), and 33.2% of MSM reported having not tested for HIV in their lifetime(3).

HIV self-testing (HIVST), recommended by the World Health Organization, may complement facility-based HIV testing and has the potential to increase HIV testing coverage(4). Self-testing involves an individual performing a test and interpreting the results themselves (WHO guidelines on self-testing). HIVST provides privacy, convenience, and confidentiality. HIVST can be used to collect blood or saliva(5). HIVST is acceptable among many HIV key populations(6). Studies conducted in China and Brazil found that HIVST could increase HIV testing frequency and increase testing coverage among MSM and their sexual partners(7, 8). However, HIVST uptake remain low in some countries, including Malaysia (25%) and Malawi (45%)(9, 10).

Social network-based strategies leverage large social networks where individuals are linked together(11). Social network-based strategies can improve HIV testing, referral, adherence, and retention among MSM(12, 13). Secondary distribution is a social network-based strategy where multiple HIV testing cards or HIVST kits are provided to individuals (referred to as “indexes”) to distribute to their sexual and social contacts (referred to as “alters”)(14). Previous studies found that the use of invitation cards during pregnancy could facilitate male partner testing and involvement in preventing mother-to-child HIV transmission(15, 16). Peer referral of HIV testing could also increase testing uptake and reach high-risk individuals(17, 18). However, most research on the distribution of testing cards required alters to test for HIV at health facilities. One study conducted in Kenya found that the provision of multiple HIVST kits to women led to higher HIV testing among their partners and higher couples testing than the distribution of invitation cards(19). In previous studies from South Africa and the U.S., secondary distribution of HIVST kits was found to be feasible and effective among MSM(20, 21). Even though there are extensive studies focused on the secondary distribution of testing cards and HIVST kits, limited data exist for the comparison of the effectiveness between these two methods among MSM. These data are helpful for informing the scale-up of HIVST in MSM. This quasi-experimental study aimed to evaluate whether the secondary distribution of HIVST kits is more effective in promoting HIV testing by comparing to distribution of testing referral cards among Chinese MSM.

## Methods

### Study design and setting

This quasi-experimental study was conducted in Guangzhou, China from May 2019 to January 2020. In this study, we compared the effectiveness of an HIVST secondary distribution model (intervention) with an HIV testing referral card distribution model (control) in promoting social network-based HIV testing. The HIVST secondary distribution program was implemented between May 1^st^ and October 7^th^ 2019 at Dermatology Hospital of Southern Medical University (DH-SMU). Study staff at the site handled the study procedures including testing, introducing the program, and offering HIVST kits to participants. The HIV testing card referral was implemented between October 14^th^ 2019 and January 17^th^ 2020 at three sites – a local MSM community-based HIV testing clinic, a local MSM community-based HIV testing weekend clinic at DH-SMU, and a municipal-level CDC HIV testing clinic. Testing card referral was embedded in their routine HIV Counseling and Testing (HCT) services. MSM volunteers or public health staff handled the same study procedures as in the intervention arm. The two arms were implemented one at a time in order to compare the intended outcomes in similar catchment areas without having people choose between the two. No significant temporal fluctuations were identified in HIV testing uptake by month. Eligibility was defined as 18 years or older, born biologically male, ever had sex with men, were willing to participate in follow-up at one month, and willing to provide a phone number.

### HIVST secondary distribution program

We posted banner ads on WeChat and Blued, the most used social media applications among Chinese population and Chinese MSM respectively. Interested men scheduled an appointment with study staff located at DH-SMU for free HIV/syphilis testing and an opportunity to apply for up to five free HIVST kits for distribution to people in their social network. At the site, indexes were given a dual HIV/syphilis test by the study staff using the same test in the HIVST kit. After indexes were introduced of the study project and how to use the HIVST in person, those who agreed to distribute HIVST to alters could sign up for up to five kits. Each kit additionally included instructions for use, a QR code for test result upload, and a unique ID number to link indexes with their alters. Indexes were requested to distribute the kits in a month and were allowed to access kits for multiple times (Figure 1).

**Figure 1.**
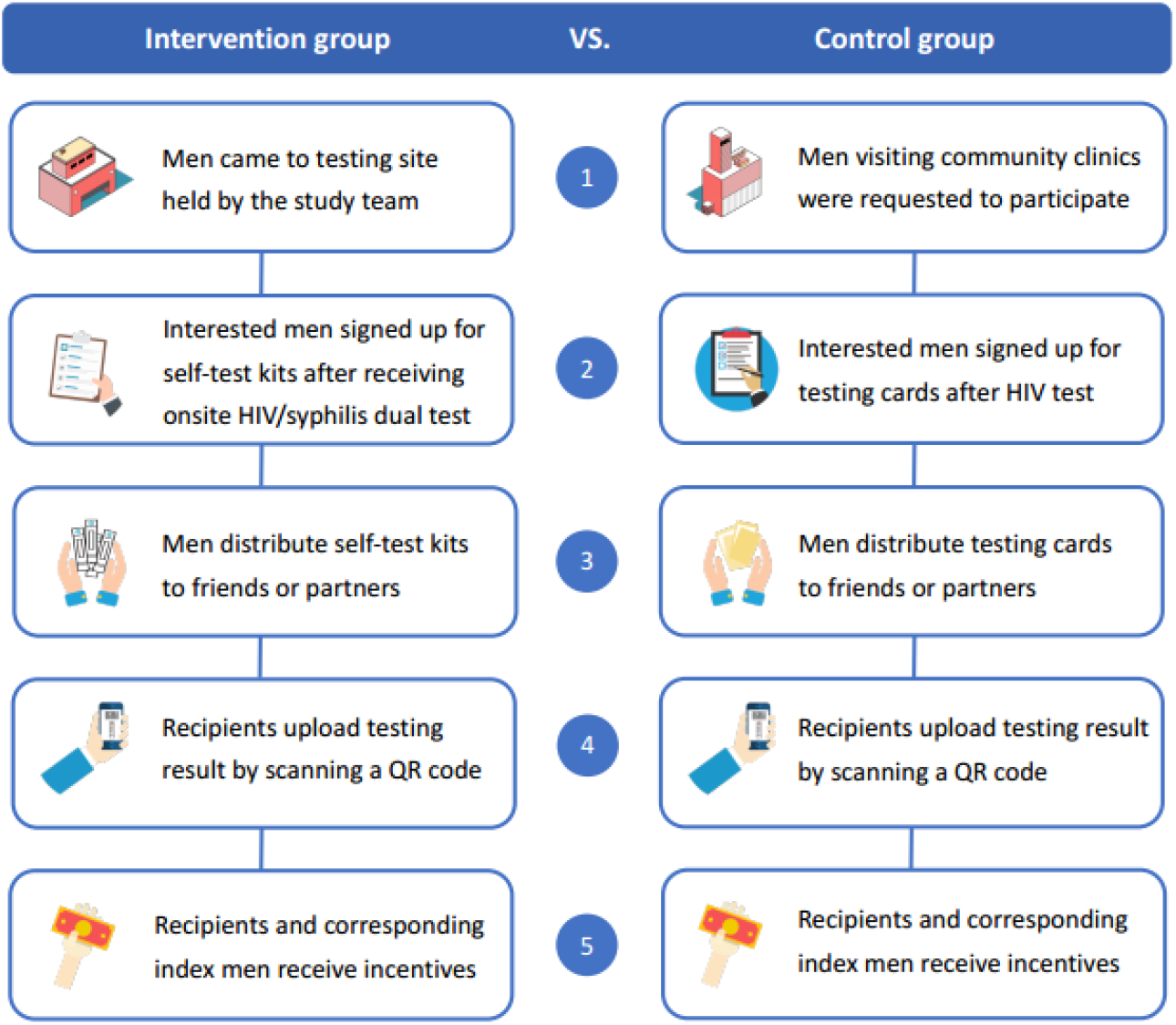
Study procedures of intervention group (secondary distribution of HIVST) and control group (HIV testing card)

After using the HIVST kit, alters were encouraged to upload their test result with the kit’s numeric ID to a database located at Sojump by scanning the QR code. Uploaded test results were verified by study staff. Alters who uploaded a reactive result were contacted to recommend confirmatory testing at a district-level CDC. All uploaded results were linked to index men. Index men received an incentive of USD$3 after completing baseline survey, and $5 for follow-up survey. Alters and corresponding index men both received an additional $3 when alters uploaded their test results.

### HIV testing card

Men visiting the three clinics for HIV testing were invited to participate after community-based volunteers or public health staff introduced the project in person at the site. Eligible men could sign up for up to five testing referral cards. Each testing card contained a QR code to upload test results when the alter completed an HIV test either in a testing facility or using a self-test, and a unique numeric ID linked alters with index men. Index men were encouraged to distribute the testing referral cards in a month.

Alters who received testing cards were encouraged to take an HIV test however they preferred. If they took a facility-based test, they were encouraged to upload a report of their test result by scanning, screenshotting, or taking a picture of the report, and uploading their test result to the study database. If they self-tested, they were encouraged to upload a picture of their self-test result. All uploads were linked to index men. Index men received an incentive of $3 after completing baseline survey, and $5 for follow-up survey. Alters and corresponding index men received an additional $3 when alters uploaded their test results.

### Data collection

We collected data from all indexes who agreed to distribute, and all alters who uploaded their test results after receiving HIVST kits/testing cards. Index men were requested to complete an online baseline survey at the site and an online follow-up survey at one month. Baseline survey items included sociodemographic information, recent sexual experiences in the past month, testing experiences, social network data and respondents’ phone number to identify men who participated more than once. Sociodemographic information included age, education, marital status, income, sexual orientation and gender identity, and disclosure as MSM. Data on sexual behaviors included previous sex with men, number of sex partners in the past month, and condomeless anal sex in the past month. Data on HIV testing behaviors included previous HIV test, previous self-test, testing locations, and comparison between self-testing and clinic-based testing. Social network data were characterized by requesting indexes to list their social contacts, describe their relationships and answer whether they would distribute to each specific contact.

A follow-up survey asked about index men’s relations to the recipients, number of HIVST kits/testing cards distributed, sexual experiences in the past month, and distributing experiences including point of sex.

Alters were also requested to complete an online survey when uploading test results. The survey instrument collected the same data as index men. Additional data on alters’ experience receiving and using HIV self-tests was gathered.

Cost data was collected for all expenditures from organizations in the two arms using a health provider perspective. Fixed costs consist of building rent, office equipment, and personnel. Variable costs include consumables, telephone bill, and transport (see supplement 1). Cost for the HIVST arm was collected from May 1^st^ to September 30^th^ 2019 over which this arm was implemented. Cost for the referral card arm was collected from October 14^st^ to January 14^th^ 2020. The primary outcomes of the analysis are the cost per alter tested and cost per alter diagnosed with HIV.

### Outcomes

The primary outcomes of this study were: 1) test uptake measured by the proportion of index men self-reporting having distributed self-test kits to sexual partners or peers; and 2) the mean number of kits distributions self-reported. Secondary outcomes were subgroup analyses based on age, sexual orientation, sexual orientation disclosure, sex in the past month, and prior HIV testing. We further compared the cost-effectiveness of HIVST secondary distribution model to testing card referral model reporting incremental cost-effectiveness ratio (ICER). We used descriptive statistics to examine sociodemographic and behavioral characteristics of index men.

### Data analysis

We examined the hypothesis that secondary distribution of HIVST would increase HIV test uptake compared to the distribution of testing card. We reported risk ratios of participants in each group who reported distributions, risk differences of the numbers of distributions participants reported, with estimated 95% confidence interval (CI) respectively. Risk ratios were calculated by unconditional maximum likelihood estimation (Wald) and were adjusted for small sample. CIs were calculated using normal approximation (Wald). We assessed effect modification using a linear probability model based on five subgroups: age (no older than 30 years old and older than 30), sexual orientation (self-identified as gay and self-identified as other than gay), disclosure (yes or no), recent sex in the past month (yes or no), and prior experience of HIV testing (ever tested for HIV or never tested for HIV). Missing data of distributions for not distributing to any type of recipients was treated as the event not detected and counted as 0. All analyses were performed in R 3.6.3.

### Ethical approval and inform consent

The study protocol was approved by the ethics review committees at the University of North Carolina at Chapel Hill and the Dermatology Hospital of Southern Medical University. Verbal consent was obtained from all participants who came to the sites and agreed to participate. Signed online consent was accessed from all index participants who completed the baseline survey and alters who submitted their testing results.

## Results

From May 2019 to January 2020, 106 index participants in the HIVST secondary distribution group and 102 index participants in the testing card referral group were recruited (Figure 2). No duplicated participant was identified across the two groups. At one month, 92 (86.8%) participants who accessed HIVST kits and 62 (60.8%) participants who accessed testing cards for distribution completed a follow-up survey. In the referral testing card arm, participants who received or opted out of follow-up were similar in sociodemographic characteristics and risk behaviors except for sexual orientation (see supplement 2).

**Figure 2.**
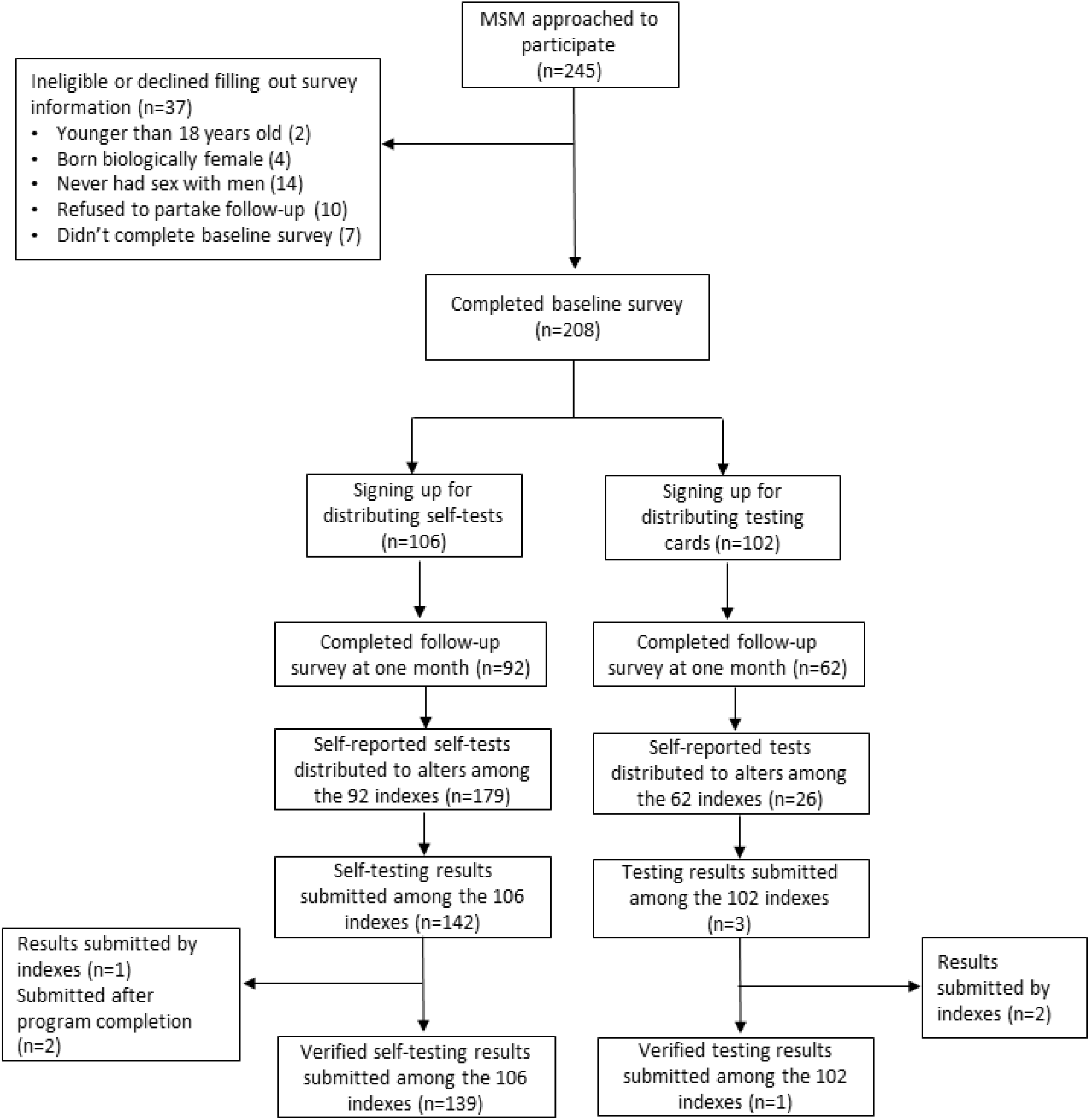
Flowchart of the study cohort.

At follow-up, 92 index participants in the HIVST arm self-reported having distributed 179 kits to alters, while indexes in the testing card arm self-reported 26 distributions. We received 142 test results uploads from the HIVST secondary distribution group, among which 139 were from unique alters, one was from an index participant, and two were submitted after the program completion. Three test results were uploaded in the testing card referral group, among them one was from a unique alter, and two were from index participants. Fourteen participants in the HIVST arm and two in the testing card arm received a reactive result. Except that one participant was confirmed to be false positive and two lost contact in the HIVST arm, the remaining participants in the study received positive confirmatory test results. We used self-reported distributions at follow-up for analysis.

### Index participants characteristics

Demographic characteristics were mostly similar between the HIVST secondary distribution group and testing card referral group (Table 1). Mean age of the participants were 27.0 years old (SD=6.74) and 28.7 years old (SD=7.33), respectively. The majority of participants were self-identified men (98%), self-identified gay (74% (HIVST) vs. 77% (Testing card)), unmarried (88% vs. 92%), had a monthly income more than USD$458 (76% vs. 82%), had a higher education degree (75% vs. 71%), and had anal sex in the past month (60% vs. 57%). A higher proportion of participants in the testing card referral arm have ever tested for HIV compared to participants in the HIVST arm (78% vs. 89%, p = 0.039).

**Table 1.** Baseline index men sociodemographic characteristics among MSM in China, 2019-2020 (N=208)

|  | Testing card (N=102) |  | HIVST (N=106) |  | P values |
| --- | --- | --- | --- | --- | --- |
| Age (mean, SD) | 28.7 | 6.74 | 27.0 | 7.33 | 0.80 |
| Gender identity (n, %) |  |  |  |  | 1.00 |
| *Men* | 100 | 98% | 104 | 98% |  |
| *Unsure* | 2 | 2% | 2 | 2% |  |
| *Women, transgender individuals* | 0 | 0% | 0 | 0% |  |
| Sexual orientation (n, %) |  |  |  |  | 0.53 |
| *Gay* | 79 | 77% | 78 | 74% |  |
| *Heterosexual* | 1 | 1% | 1 | 1% |  |
| *Bisexual* | 16 | 16% | 23 | 22% |  |
| *Unsure* | 6 | 6% | 4 | 4% |  |
| Marital status (n, %) |  |  |  |  | 0.48 |
| *Engaged or married* | 4 | 4% | 6 | 6% |  |
| *unmarried* | 94 | 92% | 94 | 88% |  |
| *divorced* | 4 | 4% | 6 | 6% |  |
| Education (n, %) |  |  |  |  | 0.61 |
| *High school or less* | 23 | 23% | 20 | 19% |  |
| *College or equivalent* | 73 | 71% | 80 | 75% |  |
| *Graduate* | 6 | 6% | 6 | 6% |  |
| Income (n, %) |  |  |  |  | 0.31 |
| *No more than \$458 per month* | 18 | 18% | 25 | 24% | |
| *More than \$458 per month* | 84 | 82% | 81 | 76% | |
| Anal in the past month (n, %) | 57 | 56% | 64 | 60% | 0.57 |
| Ever tested HIV (n, %) | 91 | 89% | 83 | 78% | 0.039 |

### Distributions from index participants to alters

More participants in the secondary distribution of HIVST kits group reported distributing at least one test (29% vs. 75%, p<0.001), at least given one test to their sexual partner (11% vs. 38%, p<0.001), and at least given one test to their friend (18% vs. 50%, p<0.001) than those in the testing card distribution group (Table 2).

**Table 2.** Proportions of index participants who reported having distributed to alters among MSM in China, 2019-2020 (N=154)

|  | Testing card<br>(N=62) | HIVST<br>(N=92) | Risk difference<br>[95% CI] | Adjusted risk ratio¶<br>[95% CI] |
| --- | --- | --- | --- | --- |
| Distributed at least one<br>test | 18 (0.29) | 69 (0.75) | 0.46 [0.31, 0.61] * | 2.49 [1.66, 3.73] * |
| Distributed at least one<br>test to sexual partner | 7 (0.11) | 35 (0.38) | 0.27 [0.14, 0.39] * | 2.99 [1.42, 6.31] * |
| Distributed at least one<br>test to friend | 11 (0.18) | 46 (0.5) | 0.32 [0.18, 0.46] * | 2.63 [1.48, 4.66] * |

¶ Small sample-adjusted UMLE & normal approx. (Wald) CI.
\* p<0.001

Participants in the HIVST secondary distribution group reported having distributed more self-tests to social contacts (0.42 vs. 1.95, p<0.001), more self-tests to sexual partner (0.21 vs. 0.63, p<0.01), and more self-tests to friends (0.23 vs. 1.13, p<0.001), compared to participants distributing testing cards(Table. 3).

**Table 3.** Numbers of index participants who reported having distributed to alters among MSM in China, 2019-2020 (N=154)

|  | Testing card (N=62) | HIVST (N=92) | Risk difference<br>[95% CI] |
| --- | --- | --- | --- |
| Average number of tests<br>distributed | 0.42 | 1.95 | 1.53 [1.09, 1.96] ** |
| Average number of tests<br>distributed to sexual partners | 0.21 | 0.63 | 0.42 [0.13, 0.71] * |
| Average number of tests<br>distributed to friends | 0.23 | 1.13 | 0.90 [0.55, 1.25] ** |

\*\* p<0.001, \* p<0.01

**Table 4.** Subgroup analyses of secondary distribution of HIVST and testing card in quasi-experimental study in China, 2019 (N=154)

| Subgroup | HIVST<br>Distributed/Total | Testing card<br>Distributed/Total | Risk difference<br>[95% CI] | P value for<br>interaction |
| --- | --- | --- | --- | --- |
| Age |  |  |  |  |
| Age ≤ 30 | 73/77 (0.95) | 10/38 (0.26) | 0.68 [0.54 to 0.83] | 0.39 |
| Age > 30 | 14/15 (0.93) | 14/22 (0.64) | 0.30 [0.06 to 0.53] |  |
| Sexual orientation |  |  |  |  |
| Gay | 68/71 (0.96) | 10/43 (0.23) | 0.73 [0.59 to 0.86] | 0.007 |
| Others | 19/22 (0.86) | 8/17 (0.47) | 0.39 [0.12 to 0.67] |  |
| Disclosure |  |  |  |  |
| Disclosed | 72/76 (0.95) | 14/48 (0.29) | 0.66 [0.52 to 0.79] | 0.72 |
| Not disclosed | 15/16 (0.94) | 4/12 (0.33) | 0.60 [0.31 to 0.90] |  |
| Anal sex in the past month |  |  |  |  |
| Yes | 41/43 (0.95) | 11/32 (0.34) | 0.61 [0.43 to 0.79] | 0.49 |
| No | 46/49 (0.94) | 7/28 (0.25) | 0.69 [0.51 to 0.86] |  |
| Experience of testing |  |  |  |  |
| Ever tested | 69/73 (0.95) | 14/54 (0.26) | 0.69 [0.56 to 0.81] | 0.02 |
| Never tested | 18/19 (0.95) | 4/6 (0.67) | 0.28 [-0.1 to 0.67] |  |

**Table 5.**
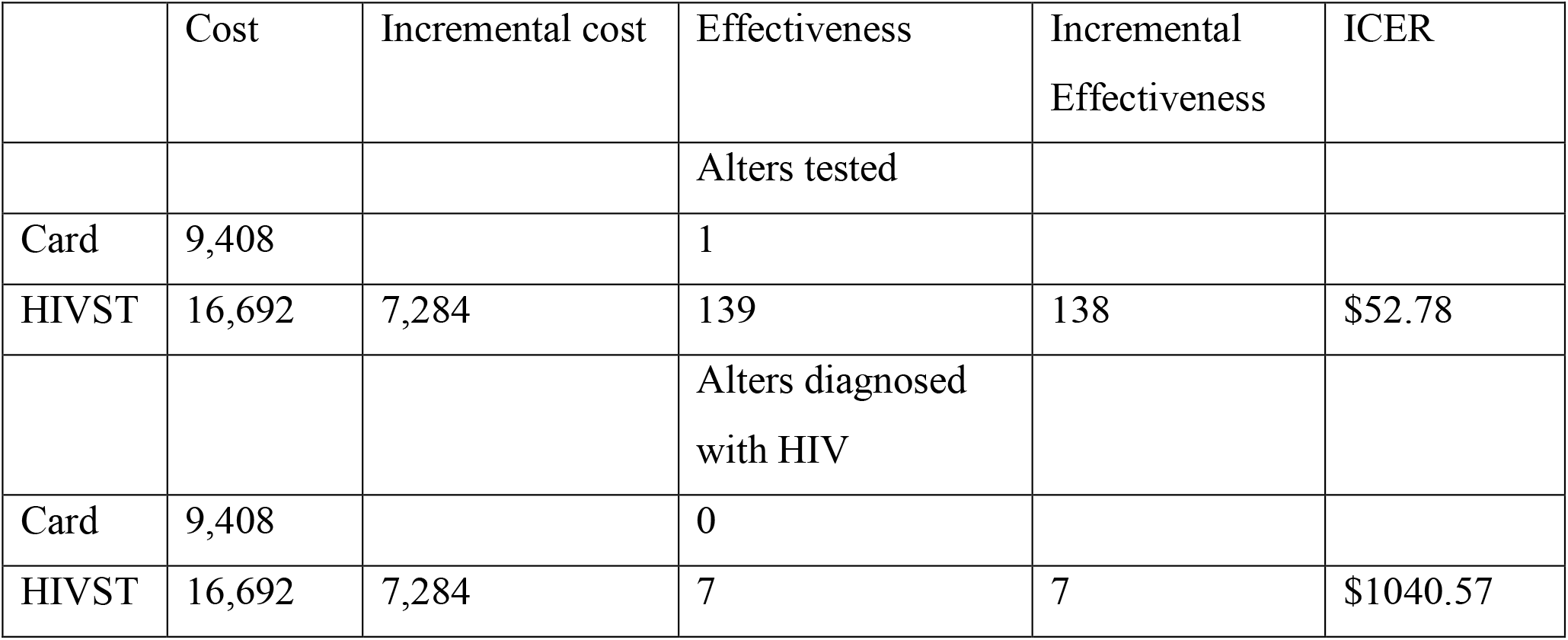
Cost-effectiveness between HIVST arm and testing card referral arm.

|  | Cost | Incremental cost | Effectiveness | Incremental Effectiveness | ICER |
| --- | --- | --- | --- | --- | --- |
|  |  |  | Alters tested |  |  |
| Card | 9,408 |  | 1 |  |  |
| HIVST | 16,692 | 7,284 | 139 | 138 | \$52.78 |
|  |  |  | Alters diagnosed with HIV |  |  |
| Card | 9,408 |  | 0 |  |  |
| HIVST | 16,692 | 7,284 | 7 | 7 | \$1040.57 |

### Distributions by subgroups

The effect of secondary distribution of HIVST is modified by self-identified sexual orientation and prior experience of testing. Participants in the intervention group who are self-identified as gay were significantly more likely to have distributing behaviors compared to participants in the same group who self-identified as other than gay or participants in the control group (p = 0.007), and participants in the intervention group who ever tested for HIV were significantly more likely to distribute compared to participants in the same group who never tested for HIV or participants in the control group (p = 0.02). There was no significant effect modification between groups of different age, different disclosure status, or different sexual behaviors in the past month.

### Costing analysis

The total economic cost for the HIVST secondary distribution group was $ 16692, and $9408 for the testing card referral distribution group, with an incremental cost of $ 7284. The cost per alter tested for the card referral group was $9408; and for the HIVST secondary distribution group was $120. The ICER per alter tested was $52.78. In addition, as 7 alters were tested positive in the HIVST secondary distribution group and none in the card referral group, the cost per alter tested positive for the HIVST secondary distribution group was $ 2384. The ICER per alter tested positive was $1040.57.

## Discussion

Promoting HIV testing among key populations is essential to reach the “95-95-95” targets of UNAIDS. We assessed the effectiveness of MSM using secondary distribution of HIVST compared to referral testing cards. Our data suggest the feasibility and effectiveness of secondary distribution of HIVST compared to testing card referral. Our findings extend existing literature by focusing on HIV social network-based testing modalities in China and conducting an economic evaluation.

Although both modalities leveraged social network to expand testing coverage, indexes who distributed HIVST were more likely to have peers receive HIV testing compared to indexes who referred HIV testing cards. Our data is consistent with prior findings suggesting HIV self-testing could circumvent barriers to HIV testing and increase testing coverage among key populations(6, 22). Compared to peer referral, secondary distribution of HIVST could not only utilize peer influence to encourage testing in a similar way as peer referral of testing(17), but also enable recipients to test in private setting with confidentiality and less stigma, which is not present in testing cards referral(23). In China, only 56.4% of Chinese MSM living with HIV were aware of their status as of 2019, while there is no available data on HIV status awareness in the general population(24). HIV self-testing and secondary distribution of HIV self-testing could be helpful to increase testing uptake among key populations in China.

We also found that participants who previously tested for HIV in the HIVST group were more likely to distribute tests to their social contacts. Retesting is recommended for key populations and in settings where there is a high proportion of people have ever tested, retesting strategies to identify new cases are needed to reach the UNAIDS target of 95% status awareness(25). Previous evidence has shown that direct provision of self-testing could facilitate retesting for HIV among key populations(26). Our data suggest that MSM who retest for HIV were also willing to distribute HIVST to their social network. Secondary distribution of HIVST could be combined with retesting strategies to increase testing uptake among MSM in China.

Secondary distribution of HIVST was cost-effective in increasing testing uptake among MSM, as the cost per alter tested in the HIVST arm was $ 120 while $9408 in the testing card referral arm. However, the total cost in the HIVST arm was higher than that in the testing card referral arm ($16692 vs. $9408), making secondary distribution of HIVST less cost-effective regarding identifying HIV cases. Although data from the US suggested that HIVST was cost-saving in terms of finding new cases compared to facility-based testing(27), economic evaluations from low- and middle-income countries (LMICs) did not show cost-savings due to relatively higher costs of HIVST kits and staff(28-30). In our study, secondary distribution of HIV self-tests was implemented by the research team in a facility-based setting where participants came to the site to test for HIV and sign up for free HIVST kits. Personnel cost comprised 59% of the total cost. Future implementation of HIVST secondary distribution should advance new modalities to reduce cost related to personnel.

This study demonstrated that embedding secondary distribution of HIVST in facility-based testing services is feasible to increase testing uptake and can be added to existing testing services in China with future implications for implementation. First, as MSM who previously tested for HIV or who self-identified as gay were more effective at distributing HIVST kits to their social network, testing facilities nationwide are recommended to provide free HIVST kits. Especially, community-based organizations should be the focal point to organize and supply HIVST to the community. Second, testing modalities that could reduce personnel costs are needed. Future approaches could include HIVST in HIV voluntary counselling and testing services and incorporate digital strategies to lower fixed cost.

Our study has several limitations. First, analyses of distributing behaviors were based on self-report. Although uploaded HIV self-test results were used to link index men who distributed HIVST kits, which enabled us to determine the actual distributions in the intervention group, the distributions in the control group were not verified due to very few alters who uploaded their test results. It could be possible that individuals overreported distributions, however, there is no reason to believe reporting between groups at follow-up would be different. Second, 13% (14/106) of index men in the intervention group and 39% (39/102) in the control group were lost to follow-up, which could introduce bias. However, those who received or opted out of follow-up were similar in sociodemographic characteristics and risk behaviors except for sexual orientation (see supplement 2).

## Conclusions

Secondary distribution of HIVST engaged more MSM to distribute tests to their social network and reached more MSM for test. MSM who self-identify as gay or who have previously tested for HIV were more effective in distributing tests. Future testing approaches should include HIVST kits in voluntary counselling and testing settings and incorporate digital strategies for secondary distribution.

**Supplement 1:** Costs related to study implementation, USD$, (1 USD = 6.54 CNY)

|  |  |
| --- | --- |
| **HIVST arm** |  |
| **Fixed cost** | **Amount** |
| Building rent | 698.01 |
| Office equipment | 107.15 |
| Personnel | 9785.93 |
| Promotion fees | 2293.58 |
| **Variable cost** |  |
| Consumables | 3777.30 |
| Telephone bill | 19.85 |
| Transport | 11.00 |
| **Testing referral card arm** |  |
| **Fixed cost** | **Amount** |
| Building rent | 1256.42 |
| Office equipment | 257.15 |
| Personnel | 6360.3 |
| **Variable cost** |  |
| Consumables | 968.40 |
| Telephone bill | 19.85 |
| Transport | 3.06 |
| Promotion fees | 542.81 |

**Supplement 2:** Comparison between participants who received and opted out of follow-up in the control group, 2019-2020, China (N=102)

|  | Followed up<br>(N=63) | Follow-up opted out<br>(N=39) | P values |
| --- | --- | --- | --- |
| Age | 29.3 (6.9) | 27.7 (6.5) | 0.52 |
| Marital status |  |  |  |
| Engaged or married | 2 (3.2%) | 2 (5.1%) | 0.64 |
| Unmarried | 57 (90.5%) | 37 (94.9%) | 0.71 |
| Divorced | 4 (6.3%) | 0 (0) | 0.30 |
| Education |  |  |  |
| High school or less | 15 (23.8%) | 8 (20.5%) | 0.81 |
| College or equivalent | 43 (68.3%) | 30 (76.9%) | 0.38 |
| Graduate | 5 (7.9%) | 1 (2.6%) | 0.40 |
| Income |  |  |  |
| Less than 3000 per month | 11 (17.5%) | 7 (17.9%) | 1 |
| More than 3000 per month | 52 (82.5%) | 32 (82.1%) |  |
| Gender identity |  |  |  |
| Men | 61 (96.8%) | 39 (100%) | 0.52 |
| Unsure | 2 (3.2%) | 0 (0) |  |
| Sexual orientation |  |  |  |
| Gay | 44 (69.8%) | 35 (89.7%) | 0.03 |
| Heterosexual | 1 | 0 | 1 |
| Bisexual | 14 | 2 | 0.03 |
| Unsure | 4 | 2 | 1 |
| Anal in the past month | 34 (54.0%) | 23 (59.0%) | 0.68 |
| Ever tested for HIV | 56 (88.9%) | 35 (89.7%) | 1 |

## Data Availability

The data are avaliable upon sending requirement to the corresponding author of the manuscript.

## Notes

### Competing Interest Statement

The authors have declared no competing interest.

### Clinical Trial

ChiCTR1900025433

### Funding Statement

This work was supported by the National Key Research and Development Program of China [grant number 2017YFE0103800]; Academy of Medical Sciences and the Newton Fund [grant number NIF\R1\181020]; the National Institutes of Health [grant numbers NIAID 1R01AI114310-01, NIAID K24AI143471, R25 AI140495]; UNC Center for AIDS Research [grant number NIAID 5P30AI050410]; National Science and Technology Major Project of China [grant number 2018ZX10101-001-001-003]; the National Nature Science Foundation of China [grant numbers 81903371, 81772240]; and Zhuhai Medical and Health Science and Technology Plan Project [grant number 20181117A010064].

### Author Declarations

The study protocol was given approval by the ethics review committees at the University of North Carolina at Chapel Hill and the Dermatology Hospital of Southern Medical University. Verbal consent was obtained from all participants who came to the sites and agreed to participate. Signed online consent was accessed from all index participants who completed the baseline survey and alters who submitted their testing results.

## References

1. Ancaids N. Update on the AIDS/STD epidemic in China in December 2017. Chin J AIDS STD. 2018;24(2):111.

2. China TNHCotPsRo. The National Health Commission of the People’s Republic of China Press Release 2018 [Available from: http://www.scio.gov.cn/m/xwfbh/gbwxwfbh/xwfbh/wsb/Document/1642083/1642083.htm.

3. Li X, Lu H, Raymond H, Sun Y, Jia Y, He X, et al. Untested and undiagnosed: barriers to HIV testing among men who have sex with men, Beijing, China. Sexually transmitted infections. 2012;88(3):187–93.

4. Zhang C, Li X, Brecht M-L, Koniak-Griffin D. Can self-testing increase HIV testing among men who have sex with men: a systematic review and meta-analysis. PLoS One. 2017;12(11):e0188890.

5. Organization WH. Guidelines on HIV self-testing and partner notification: supplement to consolidated guidelines on HIV testing services: World Health Organization; 2016.

6. Figueroa C, Johnson C, Verster A, Baggaley R. Attitudes and acceptability on HIV self - testing among key populations: a literature review. AIDS and Behavior. 2015;19(11):1949–65.

7. De Boni RB, Veloso VG, Fernandes NM, Lessa F, Corrêa RG, Lima RDS, et al. An internet-based HIV self-testing program to increase HIV testing uptake among men who have sex with men in Brazil: descriptive cross-sectional analysis. Journal of medical Internet research. 2019;21(8):e14145.

8. Zhang C, Koniak-Griffin D, Qian H-Z, Goldsamt LA, Wang H, Brecht M-L, et al. Impact of providing free HIV self-testing kits on frequency of testing among men who have sex with men and their sexual partners in China: A randomized controlled trial. PLoS medicine. 2020;17(10):e1003365.

9. Indravudh PP, Hensen B, Nzawa R, Chilongosi R, Nyirenda R, Johnson CC, et al. Who is Reached by HIV Self-Testing? Individual Factors Associated With Self-Testing Within a Community-Based Program in Rural Malawi. JAIDS Journal of Acquired Immune Deficiency Syndromes. 2020;85(2):165–73.

10. Shrestha R, Alias H, Wong LP, Altice FL, Lim SH. Using individual stated-preferences to optimize HIV self-testing service delivery among men who have sex with men (MSM) in Malaysia: results from a conjoint-based analysis. BMC Public Health. 2020;20(1):1–11.

11. McCree DH, Millett G, Baytop C, Royal S, Ellen J, Halkitis PN, et al. Lessons learned from use of social network strategy in HIV testing programs targeting African American men who have sex with men. American journal of public health. 2013;103(10):1851–6.

12. Ghosh D, Krishnan A, Gibson B, Brown S-E, Latkin CA, Altice FL. Social network strategies to address HIV prevention and treatment continuum of care among at-risk and HIV-infected substance users: a systematic scoping review. AIDS and Behavior. 2017;21(4):1183–207.

13. Wang K, Brown K, Shen SY, Tucker J. Social network-based interventions to promote condom use: a systematic review. AIDS Behav. 2011;15(7):1298–308.

14. Thirumurthy H, Masters SH, Mavedzenge SN, Maman S, Omanga E, Agot K. Promoting male partner HIV testing and safer sexual decision making through secondary distribution of self-tests by HIV-negative female sex workers and women receiving antenatal and post-partum care in Kenya: a cohort study. The Lancet HIV. 2016;3(6):e266–e74.

15. Nyondo AL, Choko AT, Chimwaza AF, Muula AS. Invitation cards during pregnancy enhance male partner involvement in prevention of mother to child transmission (PMTCT) of human immunodeficiency virus (HIV) in Blantyre, Malawi: a randomized controlled open label trial. PLoS One. 2015;10(3):e0119273.

16. Koo K, Makin JD, Forsyth BW. Where are the men? Targeting male partners in preventing mother-to-child HIV transmission. AIDS care. 2013;25(1):43–8.

17. Ong’wen P, Samba BO, Moghadassi M, Okoko N, Bukusi EA, Cohen CR, et al. Chain Peer Referral Approach for HIV Testing Among Adolescents in Kisumu County, Kenya. AIDS Behav. 2020;24(2):484–90.

18. Glasman LR, Dickson-Gomez J, Lechuga J, Tarima S, Bodnar G, de Mendoza LR. Using Peer-Referral Chains with Incentives to Promote HIV Testing and Identify Undiagnosed HIV Infections Among Crack Users in San Salvador. AIDS Behav. 2016;20(6):123 6–43.

19. Masters SH, Agot K, Obonyo B, Mavedzenge SN, Maman S, Thirumurthy H. Promoting partner testing and couples testing through secondary distribution of HIV self-tests: a randomized clinical trial. PLoS medicine. 2016;13(11).

20. Lightfoot MA, Campbell CK, Moss N, Treves-Kagan S, Agnew E, Dufour M-SK, et al. Using a social network strategy to distribute HIV self-test kits to African American and Latino MSM. JAIDS Journal of Acquired Immune Deficiency Syndromes. 2018;79(1):38–45.

21. Lippman SA, Lane T, Rabede O, Gilmore H, Chen Y-H, Mlotshwa N, et al. High acceptability and increased HIV testing frequency following introduction of HIV self-testing and network distribution among South African MSM. Journal of acquired immune deficiency syndromes (1999). 2018;77(3):279.

22. Johnson CC, Kennedy C, Fonner V, Siegfried N, Figueroa C, Dalal S, et al. Examining the effects of HIV self-testing compared to standard HIV testing services: a systematic review and meta-analysis. Journal of the International AIDS Society. 2017;20(1):21594.

23. den Daas C, Geerken MBR, Bal M, de Wit J, Spijker R, Op de Coul ELM. Reducing health disparities: key factors for successful implementation of social network testing with HIV self-tests among men who have sex with men with a non-western migration background in the Netherlands. AIDS Care. 2020;32(1):50–6.

24. UNAIDS. HIV testing and status awareness among men who have sex with men 2019 [Available from: https://aidsinfo.unaids.org/.

25. Chamie G, Napierala S, Agot K, Thirumurthy H. HIV testing approaches to reach the first UNAIDS 95% target in sub-Saharan Africa. The lancet HIV. 2021;8(4):e225–e36.

26. Ortblad K, Kibuuka Musoke D, Ngabirano T, Nakitende A, Magoola J, Kayiira P, et al. Direct provision versus facility collection of HIV self-tests among female sex workers in Uganda: A cluster-randomized controlled health systems trial. PLoS Med. 2017;14(11):e1002458.

27. Shrestha RK, Chavez PR, Noble M, Sansom SL, Sullivan PS, Mermin JH, et al. Estimating the costs and cost-effectiveness of HIV self-testing among men who have sex with men, United States. Journal of the International AIDS Society. 2020;23(1):e25445.

28. Cambiano V, Ford D, Mabugu T, Napierala Mavedzenge S, Miners A, Mugurungi O, et al. Assessment of the Potential Impact and Cost-effectiveness of Self-Testing for HIV in Low-Income Countries. The Journal of infectious diseases. 2015;212(4):570–7.

29. George G, Chetty T, Strauss M, Inoti S, Kinyanjui S, Mwai E, et al. Costing analysis of an SMS-based intervention to promote HIV self-testing amongst truckers and sex workers in Kenya. PLoS One. 2018;13(7):e0197305.

30. Mangenah C, Mwenge L, Sande L, Ahmed N, d’Elbée M, Chiwawa P, et al. Economic cost analysis of door-to-door community-based distribution of HIV self-test kits in Malawi, Zambia and Zimbabwe. Journal of the International AIDS Society. 2019;22 Suppl 1(Suppl Suppl 1):e25255.

